# Assessment of a Novel Method for Non-invasive Sampling of the Distal Airspace in Acute Respiratory Distress Syndrome Patients Receiving Inhaled Sedation with Sevoflurane: the ANAISS Study Protocol

**DOI:** 10.1101/2020.10.05.20207217

**Authors:** Pierre-Antoine Tronche, Robin Lalande, Raiko Blondonnet, Laurence Roszyk, Ruoyang Zhai, Dominique Morand, Bruno Pereira, Vincent Sapin, Jean-Marc Malinovsky, Bruno Mourvillier, Jean-Michel Constantin, Joël Cousson, Matthieu Jabaudon

**Author notes:** Corresponding author and address for reprints: Matthieu Jabaudon, M.D., Ph.D., Department of Perioperative Medicine, CHU Clermont-Ferrand GReD, CNRS, INSERM, Université Clermont Auvergne, Division of Allergy, Pulmonary and Critical Care Medicine, Vanderbilt University Medical Center, Nashville, USA, 1 Place Lucie Aubrac, 63003 Clermont-Ferrand Cedex 1, France, (Mail).

## Abstract

**Introduction:** Recently, fluid collected from the heat-and-moisture-exchange filters, which are commonly used in most mechanically ventilated patients under intravenous sedation, has been reported as a potential surrogate for fluid in the distal airspace. Therefore, collection of this fluid represents a promising, non-invasive method for sampling the distal airspace in patients with acute respiratory distress syndrome (ARDS) and for facilitating a mechanistic understanding of this devastating disease. The current study protocol was constructed to assess whether this fluid could be sampled from a dedicated device (Anaesthetic Conserving Device [AnaConDa-S], Sedana Medical, Danderyd, Sweden) used to deliver inhaled sevoflurane for sedation in patients with ARDS.

**Methods and analysis:** A total of 30 adult patients within 24 hours of meeting the Berlin criteria for moderate-severe ARDS and receiving inhaled sevoflurane as standard sedation in participating centres will be eligible for inclusion into this investigator-initiated, exploratory, prospective, bicentre study. After at least 12 h of inhaled sedation, a sample of directly aspirated, undiluted pulmonary oedema fluid will be collected concurrently with fluid from the AnaConDa-S device. Levels of proinflammatory cytokines (IL-1β, IL-6, IL-8, TNF-α and sTNFr-1) and markers of lung endothelial (Ang-2) and epithelial (sRAGE) injury will be measured in both fluids by Multiplex. The primary endpoint is the correlation between protein markers (IL-1β, IL-6, IL-8, TNF-α, sTNFr-1, Ang-2 and sRAGE) measured in the undiluted pulmonary oedema fluid versus the AnaConDa-S fluid.

**Ethics and dissemination:** The study was approved by the appropriate ethics committee (*CPP Est I*). Informed consent is required. The fluid collection from the AnaConDa-S has potential to foster our understanding of the potential effects of inhaled sedation in clinical ARDS and to open up novel perspectives for prognostic and predictive enrichment in future trials. The results will be published in a peer-reviewed journal.

**Registration number:** NCT03964155.

## INTRODUCTION

### Background and rationale

Acute respiratory distress syndrome (ARDS) continues to be a major critical care concern, with high mortality rates and long-term impact in survivors (1,2). These issues are particularly highlighted by the current coronavirus disease 2019 (COVID-19) outbreak (3). No targeted pharmacologic therapies are yet available to treat this syndrome, and its management is currently based only on the treatment of its cause(s), intensive care unit (ICU) supportive care, lung-protective mechanical ventilation, and conservative fluid management whenever possible (1,2). A major limitation impeding the development of new therapies for ARDS is the difficulty in obtaining an easily reproducible and comparable sample from the injured distal airspace. Each of the currently available methods for studying the airspace of ARDS patients has significant drawbacks. For example, bronchoalveolar lavage (BAL) can be difficult to perform, is impractical for serial collections and can introduce an unknown dilution factor to the measured analytes (4). The direct aspiration of undiluted pulmonary oedema fluid (OF) is possible (5–8), but this is feasible only early in the disease course and in patients with a significant degree of oedema. Exhaled breath condensate may be useful for characterizing the airspace (9), but the current collection equipment is costly, the sample volume is limited, and the collection is labour-intensive. Identification of a simple, non-invasive way to accurately sample the distal airspace in ARDS would be a major advance for studying the biology of injured lungs.

The heat-and-moisture-exchanger (HME) filter is an in-line, disposable, hygroscopic bacteriostatic sponge that is routinely placed between the patient and the ventilator to minimize the loss of heat and moisture from exhaled breath (10,11). During mechanical ventilation, moisture from the patient’s exhaled breath condenses on this filter until the filter is exchanged after several hours. A wide variety of HME filter devices are commercially available, with varying filter designs and dead space volumes. HME filters are produced for neonatal and pediatric use as well. Very recently, fluid collected from HME filters used in most mechanically ventilated patients under intravenous sedation has been reported as a potential surrogate for fluid in the distal airspace, thereby representing a promising, simple and non-invasive method to sample the distal airspace in patients with ARDS. This sampling is also a powerful tool to facilitate mechanistic understanding of this syndrome (12).

Inhaled sedation with halogenated agents, such as isoflurane or sevoflurane, is now available to critical physicians worldwide using disposable medical devices such as the AnaConDa-S (Sedana Medical, Danderyd, Sweden) (13–15). The AnaConDa-S also acts as a HME filter and contains a bacterial/viral filter. During breathing, the volatile anesthetic agent is re-circulated through the reflector (consisting of an active carbon filter) that is also included in the device. The first pilot randomized controlled trial in patients with moderate-severe ARDS confirmed that inhaled sevoflurane improved arterial oxygenation compared to intravenous midazolam, while decreasing plasma and alveolar marker levels of inflammation (interleukin [IL]-1β, IL-6, IL-8 and tumour necrosis factor [TNF]-α) and lung epithelial injury (soluble receptor for advanced glycation end-products [sRAGE]) (16). These results, along with preclinical evidence (17–20), suggest that inhaled sevoflurane may have lung-protective effects in ARDS, although the mechanism warrants further investigation (13,21). Taken together, the available clinical evidence supports the safe use of inhaled sevoflurane for ICU sedation, with no major adverse effect on processes such as renal function or respiratory mechanics (22–25). It is now used as standard practice in some French ICUs (26).

Because we hypothesised that the condensed fluid from the AnaConDa-S device could be representative of distal airspace fluid in patients receiving inhaled sevoflurane, the *Assessment of a Novel Method for Non-invasive Sampling of the Distal Airspace in Acute Respiratory Distress Syndrome Patients Receiving Inhaled Sedation with Sevoflurane* (ANAISS) study protocol was designed to assess whether the composition of the AnaConDa-S fluid could represent that of the distal airspace fluid in patients with ARDS.

### Objectives

#### Primary objective

The primary objective is to compare the protein composition of simultaneously collected undiluted pulmonary OF and AnaConDa-S fluid from patients with moderate-to-severe ARDS.

#### Secondary objectives

A further aim of the study will be to evaluate the correlation between protein marker levels measured in AnaConDa-S and the Radiographic Assessment of Lung Edema (RALE) score (27,28); to compare protein marker levels measured in AnaConDa-S fluid between lung imaging phenotypes of focal and nonfocal ARDS (29–31); to compare protein marker levels in undiluted OF and in plasma; and to assemble a biological collection of AnaConDa-S fluids for future mechanistic studies of inhaled sevoflurane in ARDS.

### Study design

The ANAISS study is an investigator-initiated, prospective, bicentre, observational study with a collection of biological samples that will include invasive alveolar (OF) sampling. Therefore, it is labelled as a “Research Involving the Human Person (RIHP) Category 2 protocol” according to French regulations (Jardé law 2012-300, voted March 2012).

Because inhaled sedation with sevoflurane is current standard practice in participating ICUs, the study protocol includes no other specific intervention apart from the alveolar and AnaConDa-S sampling.

## METHODS: PARTICIPANTS, INTERVENTIONS AND OUTCOMES

### Study setting

The ANAISS study involves a total of two French centres (two medical-surgical ICUs at CHU Clermont-Ferrand and one medical ICU at CHU Reims) and academic research partners at Université Clermont Auvergne (Clermont-Ferrand, France) and Vanderbilt University Medical Center (Nashville, USA).

### Eligibility criteria

#### Inclusion criteria

All adult patients within 24 h from meeting the Berlin criteria for moderate or severe ARDS (32) and receiving inhaled sedation with sevoflurane as standard practice will be eligible for inclusion in the study.

#### Exclusion criteria

Patients under 18 years old and those declining to provide informed consent will not be included in the study.

### Interventions

In this study, no specific interventions will be performed apart from a single sampling of lung OF, AnaConDa-S fluid, and blood.

Patients within 24 h of moderate-to-severe ARDS onset and receiving inhaled sedation with sevoflurane as current practice in participating centres will be enrolled. Overall intensive care management practice in both participating ICUs will not be influenced by the study protocol.

After at least 12 h of inhaled sedation with sevoflurane through the AnaConDa-S device, a sample of directly-aspirated, undiluted OF will be collected in a 4ml-EDTA tube, as previously described (5–8). Concurrently, fluid will be collected from the AnaConDa-S device (removed and changed for the first time simultaneously with pulmonary sampling, and then routinely removed at least every 24 h, as per manufacturer instructions).

The AnaConDa-S device will be closed at each end with specific plugs and further processed under a laboratory hood (according to local safety policies) by suctioning the contents of the device through a BAL vial connected between the AnaConDa-S and a vacuum system, at maximal depression (inducing air intake with the thumb at the other end of the device, if needed) (**Figure**). The collected fluid will be pipetted into a 4 mL-EDTA tube and centrifuged at 2,000 × g for 10 min. The supernatant will be evenly distributed into two cryotubes stored at -80°C at each local laboratory for further analysis. Undiluted pulmonary OF will be centrifuged at 2,000 × g for 10 min; the supernatant will be distributed into two cryotubes stored at -80°C. A 4 mL blood sample will be simultaneously collected from an indwelling catheter (routinely used in ICU patients) into an EDTA tube, centrifuged at 2,000

**Figure.**
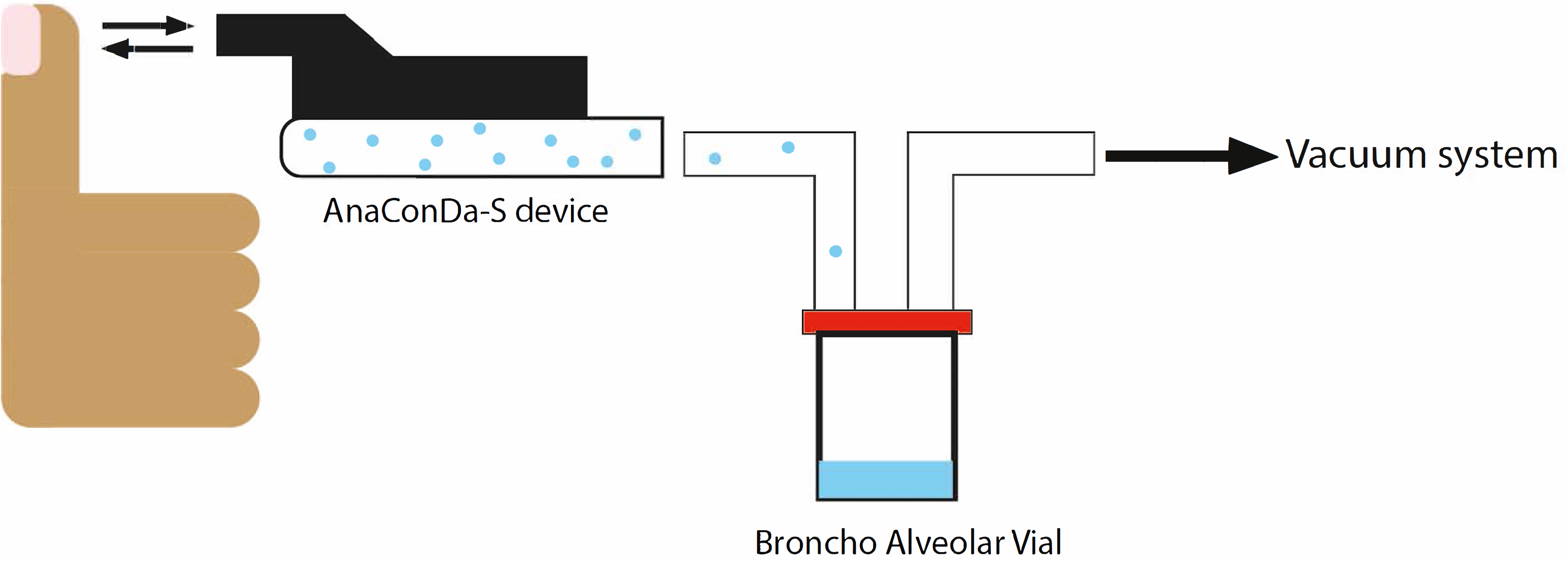
Vacuum aspiration technique used to collect fluid from the AnaConDa-S device.

× g for 10 min, and the supernatant (plasma) aliquoted and stored into a cryotube at -80°C. At the end of the recruitment period, samples will be transferred from laboratories at each site to the Department of Medical Biochemistry and Molecular Genetics at CHU Clermont-Ferrand for further analysis.

Because of the current health crisis caused by the emergence of the SARS-CoV2 virus, specific protective measures must be followed for the collection of samples if the patient is positive or considered at risk. The patient will have to be hospitalised in a dedicated specialized COVID unit, in an individual room with negative pressure with air treatment, or failing this, at neutral pressure with closed doors and sufficient ventilation. The protections for the sampler in the room will follow the protections necessary for an air and contact isolation (hat covering hair and ears, goggles, FFP2 mask, long-sleeved waterproof overshirt, gloves with a strict dressing / undressing procedure) because of the important risk of aerosolisation of the procedure. OF will be sampled in a BAL vial with a closed-suction system to avoid lung derecruitment and transferred into 4ml-EDTA tube, placed immediately in a hermetically-sealed triple packaging. Filter fluid extraction will be performed directly in the room using the ICU vacuum system, following the same instructions than for OF.

Centrifugation procedures must be carried out in a biosafety level 2 laboratory under class II biological safety cabinet, following safety guidance for high-risk sample management in patients with COVID-19 (33,34).

Levels of IL-1β, IL-6, IL-8, TNF-α, sTNFr-1, Ang-2 and sRAGE will be measured in duplicate using Multiplex (BioRad, Hercules, CA, USA) in all samples. These markers were selected a priori because they have previously been reported as markers of a hyperinflammatory ARDS phenotype (IL-1β, IL-8, IL-6, TNF-α, sTNFr-1) (35–38), of ventilator-induced lung injury (IL-6) (39), and of lung endothelial (Ang-2) (40,41) and epithelial (sRAGE) (7,42) injury in ARDS.

### Outcomes

#### Primary outcome measures

The primary outcome measure is the correlation between protein marker levels measured in the undiluted pulmonary OF and those measured in the AnaConDa-S fluid.

#### Secondary outcome measures

Secondary outcome measures are: the correlation between protein marker levels (IL-1*β*, IL-6, IL-8, TNF-*α*, sTNFr-1, Ang-2 and sRAGE) measured in the AnaConDa-S fluid and the RALE score, the difference in protein marker levels measured in the AnaConDa-S fluid between patients with focal versus nonfocal ARDS, the correlation between protein marker levels measured in undiluted OF versus plasma, and the correlation between protein marker levels measured in the AnaConDa-S fluid versus the plasma.

### Participant timeline

The participant timeline is described in the **Table**.

**Table.**
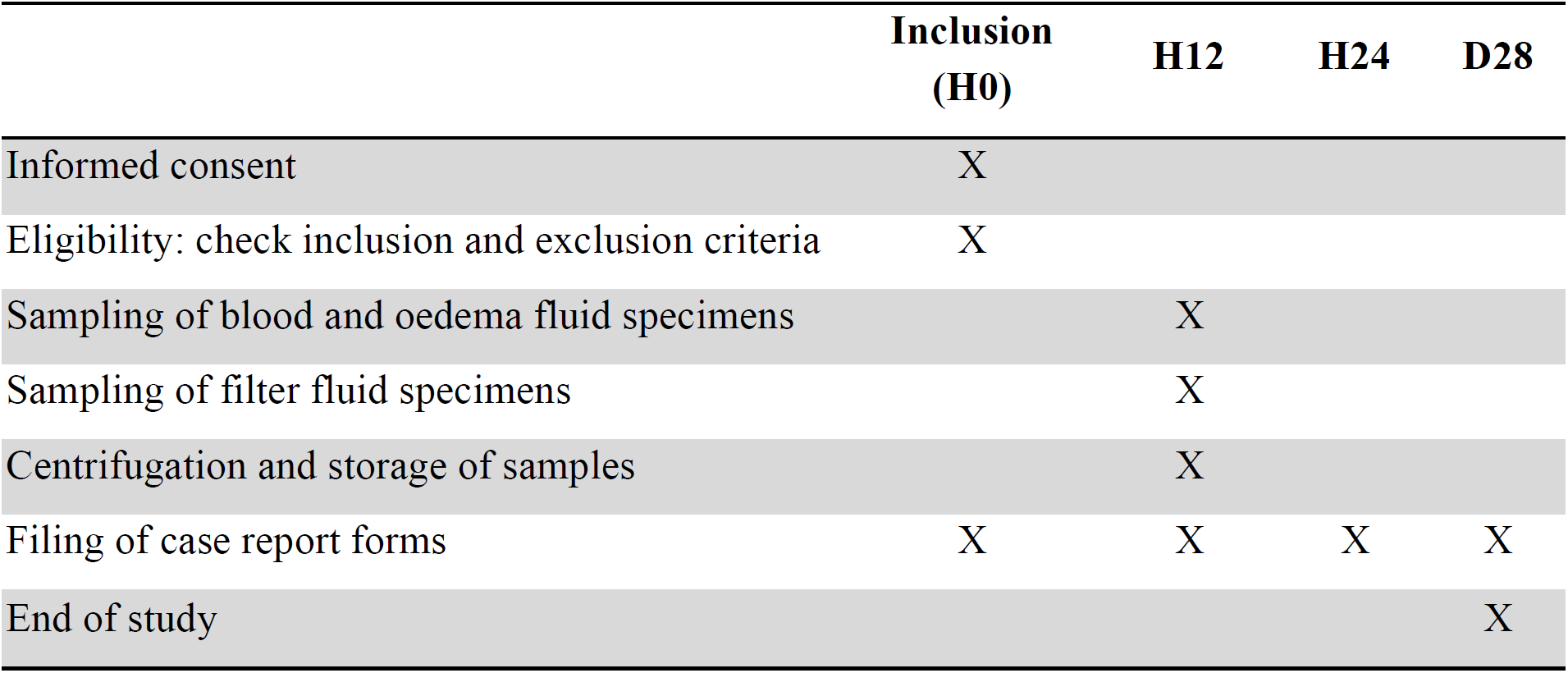
Participant timeline.

|  | **Inclusion**<br>**(H0)** | **H12** | **H24** | **D28** |
| --- | --- | --- | --- | --- |
| Informed consent | X |  |  |  |
| Eligibility: check inclusion and exclusion criteria | X |  |  |  |
| Sampling of blood and oedema fluid specimens |  | X |  |  |
| Sampling of filter fluid specimens |  | X |  |  |
| Centrifugation and storage of samples |  | X |  |  |
| Filing of case report forms | X | X | X | X |
| End of study |  |  |  | X |

### Recruitment

Patient recruitment is expected to proceed during an 18-month inclusion period beginning in September 2020.

This duration was estimated based on a high incidence of ARDS in the ICU setting (10% of ICU admissions, and nearly 25% of ICU patients requiring mechanical ventilation, in a large recent observational study) (43) and available data from each participating centre that indicate the study is highly feasible (each of the two centres will need to include less than one patient per month, holidays excluded).

2018–2020: Writing of the protocol, approvals from the ethics committee (*CPP Est I*) and the French drug agency (*Agence Nationale de Sécurité du Médicament*, ANSM); study tool development (electronic case report form, standard operating procedures for biological sampling and processing).

2020–2022: Inclusion of patients.

2022: Clean-up and closure of the database. Measurements of protein markers, data analyses, writing of the manuscript and submission for publication.

An extension of the timeline will be considered, if needed, based on the observed rate of inclusion during the recruitment period.

## METHODS: DATA COLLECTION, MANAGEMENT AND ANALYSIS

### Data collection and management

The study data will be prospectively collected and managed by trained research coordinators and/or investigators at each participating centre, using the REDCap electronic data capture tools hosted at CHU Clermont-Ferrand (44). REDCap (Research Electronic Data Capture) is a secure, web-based application designed to support data capture for research studies. It provides an intuitive interface for validated data entry, audit trails for tracking data manipulation and export procedures, automated export procedures for seamless data downloads to common statistical packages, and procedures for importing data from external sources. Data registration will be monitored by trained research coordinators.

Patient data prospectively collected from the medical record will include demographics, ARDS risk factor(s), Murray’s lung injury score (45), Acute Physiology and Chronic Health Evaluation (APACHE) II score (46) and chest x-ray at study enrolment. The ventilator settings at the time of biological sampling, the hospital and ICU length of stays, and the hospital mortality will also be recorded.

### Statistical methods

#### Sample size estimation

This study has been primarily designed as an exploratory proof-of-concept pilot study; therefore, performing a formal sample size calculation is difficult. To date, only one study (including nearly 30 patients with ARDS) has reported that HME filter fluid is representative of distal airspace fluid (12). However, in order to highlight a concordance coefficient greater than 0.7 with a two-sided type I error at 0.001 (due to multiple tests) and a statistical power around 90%, we have decided to enrol 30 patients in the ANAISS study.

An exploratory sequential analysis is planned at every 10 inclusions, without correction of type I error, to estimate the statistical power according to effect size (here, the correlation coefficient) and 95% confidence interval.

#### Statistical analysis

All analyses will be performed with Stata software (version 15, StataCorp, College Station, USA). A two-sided P value of less than 0.05 will be considered for statistical significance. However, when appropriate, a correction of the type I error (Sidak’s inflate) could be performed for multiple comparisons. Categorical data will be presented as exact numbers and percentages, whereas continuous variables will be presented as means and standard-deviations or as medians and interquartile ranges, according to their statistical distributions. The Shapiro-Wilk test will be used to assess normality.

##### Primary analysis

The concordance between marker levels measured in the undiluted OF versus AnaConDa-S fluid will be assessed using Lin’s concordance coefficient, a correlation coefficient (Pearson or Spearman, according to statistical distribution), and Bland-Altman plotting. The 95% confidence intervals will be reported. Passing and Bablok regression will be used to evaluate non-systematic and non-proportional differences in the agreement between the two sample types (undiluted OF and AnaConDa-S fluid).

##### Secondary analyses

Protein marker levels measured in the AnaConDa-S fluid will be compared between patients with focal versus nonfocal ARDS using the unpaired t-test or the Mann-Whitney U-test when appropriate. The Shapiro-Wilk and the Fisher-Snedecor tests will be used to assess normality and homoscedasticity, respectively. The receiver operating characteristic (ROC) curve will then be computed and the area under the curve will be used to evaluate which protein marker levels, as measured in the AnaConDa-S fluid, distinguish focal versus nonfocal ARDS.

The concordance between marker levels measured in the undiluted OF versus the plasma, and between those measured in the AnaConDa-S fluid versus the plasma, will be explored in a pairwise manner using Lin’s concordance coefficient, a correlation coefficient (Pearson or Spearman according to the statistical distribution), and Bland-Altman plotting. The 95% confidence intervals will be reported. Passing and Bablok regression will be used to evaluate non-systematic and non-proportional differences between sampling methods (plasma versus OF or AnaConDa-S fluid)

For each protein marker, areas under the ROC curves will be reported with 95% confidence intervals and will be compared with those of other biomarkers (47,48). Following the suggestion of Ray et al., the value of a biomarker will be considered relevant when the lower bound of the confidence interval of the area under the ROC curve is >0.75 (49).

The relationship between the protein marker levels measured in the AnaConDa-S fluid and the RALE score will be studied using correlation coefficients. Pearson’s correlation analysis will be used for variables that follow a normal distribution, and Spearman’s rank correlation coefficient analysis for variables that do not.

##### Method for missing data

No missing data are expected (few data to collect, few patients to enrol within 2 experienced centres). However, a sensitivity analysis will be performed in order to study the nature of missing data (missing at random or not) and to apply the most appropriate approach to the imputation of missing data.

##### Blinding

In the ANAISS study, the sampling method will be unblinded both to patients (if not under deep sedation) and to clinical staff members. However, to reduce the risk of bias, the investigators and personnel in charge of measuring protein markers will be unaware of the fluid type (either AnaConDa-S fluid or undiluted OF). The study statistician will also be blinded during statistical analyses.

## ETHICS AND DISSEMINATION

### Research ethics approval

The ANAISS study will be conducted in accordance with the Declaration of Helsinki and has been registered at http://www.clinicaltrial.gov on May 28, 2019 with identification number NCT03964155. The study protocol was approved by the ethics committee *CPP Est I* and the ANSM in January 2019 (approval numbers RBHP 2018 JABAUDON 2 - ANAISS 2018-A02596-49). If any further changes arise in the eligibility criteria, outcome evaluations, and/or the analysis plan, these would be submitted for approval from the ethics committee and the ANSM.

### Consent or assent

Because patients will be receiving inhaled sedation, they will not have full capacity to provide informed consent for enrolment in the study; therefore, the ANAISS study protocol provides for a waiver of informed consent from the patient.

In addition, in case consent from the patient’s next of kin cannot be obtained, local investigators will be able to include the patient in the study using an emergency consent procedure. In this case, no consent from the patient’s next of kin will be required, but the investigator will inform the patient’s next of kin of the decision as soon as possible. As per French law and good research practice, deferred informed (oral or written) consent from the patient will be sought as soon as technically possible for potential continuation of the research.

### Confidentiality

As per the French Public Health Code (articles L.1121-3 and R.5121-13), research investigators and staff members with direct access to data (who are all bound by professional secrecy, in accordance with articles 226-13 and 226-14 from the French Penal Code) shall take all necessary precautions to ensure patient data confidentiality.

In particular, all investigators and staff members shall ensure anonymity for all data collected and transferred (through the electronic case report form) to the study sponsor (CHU Clermont-Ferrand) by utilising a study-specific, anonymous identifying number. The sponsor will ensure that each research subject has given consent for study participation and data collection. Data will be handled in a confidential and anonymous manner by the sponsor, according to French law. All original records will be archived at trial sites for 15 years, as will the clean anonymised database file.

### Funding

No specific funding has been received for this study apart from institutional funds.

### Dissemination policy

Findings will be published in peer-reviewed journals and presented at local, national, and international meetings and conferences to publicise and explain the research to clinicians, commissioners, and service users. All investigators will have access to the final dataset. Both study sites will be acknowledged, and all investigators at these sites will appear with their names under ‘the ANAISS investigators’ in an Appendix to the final study report. Authorship will be granted according to the Vancouver definitions. Funding sources will have no influence on data handling, analysis or writing of the manuscript. Side studies will be allowed if approved by the scientific committee (RB, VS, LBW, JAB, JMC, and MJ), and anonymised participant-level data sets will be made accessible on a controlled-access basis.

## DISCUSSION

The primary hypothesis of the ANAISS study is that fluid collection from the AnaConDa-S device could be a novel method for assessing the distal airspace in mechanically ventilated patients with ARDS who are receiving inhaled sedation with sevoflurane. If successful, this study will have potential to further inform our understanding of the effects of inhaled sevoflurane on lung injury during ARDS through the use of a non-invasive, inexpensive and simple method.

Biomarker studies may have important value by providing a better understanding of biological mechanisms, for assessing severity and prognosis, for identifying distinct biological or radiographic phenotypes, and for predicting and/or monitoring response to therapy in ARDS (7,37,50–52). From this perspective, assessing the composition of the AnaConDa-S fluid, while validating previous findings with HME filters (12), might open up new perspectives for prognostic and predictive enrichment of future clinical trials of inhaled sevoflurane for sedation in patients with ARDS.

This study has several limitations. One limitation is the use of inhaled sevoflurane for ICU sedation, such as with the AnaConDa-S device, is not possible in all countries worldwide and remains rare overall in countries where available (26). Although this method of sedation has been reported as safe in the literature and can be considered as first-line sedation in some countries (53), its impact on major patient outcomes, such as mortality, occurrence of delirium, and post-intensive care syndrome, deserves further investigation. Several clinical trials are currently recruiting patients to address these effects of inhaled sevoflurane or isoflurane, such as the *SEvoflurane for Sedation in ARds* (SESAR, ClinicalTrials.gov Identifier: NCT04235608) or the *Comparison of an Inhaled Sedation Strategy to an Intravenous Sedation Strategy in Intensive Care Unit Patients Treated With Invasive Mechanical Ventilation* (INASED, ClinicalTrials.gov Identifier: NCT04341350) trials. The findings from this pilot ANAISS study, if positive, would enable further application of a simple method for phenotyping patients with ARDS using non-invasive sampling of the distal airspace and investigating biomarker-based enrichment methods in future research. A second limitation is the difficulty in sample size estimation, as this is a pilot, exploratory physiological study. However, with the recruitment of 30 patients, a power of 90% will be ensured to detect concordance coefficients >0.7 between protein marker levels in the undiluted OF and the AnaConDa-S fluid, with a two-sided type I error set at 0.001 due to multiple tests. A further limitation is that the method used to collect the AnaConDa-S fluid may be associated with some technical issues. In preliminary tests, we found that the suction technique described in this protocol will be the most likely to enable fluid collection from the AnaConDa-S device in an efficient and reproducible manner. However, this method requires specific equipment and strict adherence to the adequate safety policies on airway sample processing, particularly in the current context of SARS-COV2 epidemic. In addition to these technical considerations, the only risks for the patient related to the ANAISS study protocol may be those related to the sampling method for the undiluted OF. However, these risks are minor, infrequent, and reversible in most cases (e.g. alveolar de-recruitment, transient and moderate decreases in arterial oxygenation), and both participating centres have safe and strong experience in this technique (6–8,54).

This study also has several strengths. To our knowledge, it is the first study to focus on a non-invasive method of collecting biomarkers from ARDS patients receiving inhaled sevoflurane for sedation. In the future, this type of method may allow regular monitoring of pathophysiology and responses to therapeutic interventions in patients with ARDS. The assembly of a biobank of biological (alveolar, plasma, and AnaConDa-S fluid) samples will also allow future translational studies that will ultimately further our understanding of the lung-protective effects of sevoflurane (13,21). Finally, although study samples will be collected in an open-label manner, all biological measurements and statistical analyses will be performed in a blinded approach to limit the risk of evaluation bias.

In conclusion, the ANAISS study is an investigator-initiated, prospective, bicentre study that was primarily designed to assess the concordance in biomarker measurements between undiluted lung oedema fluid and fluid collected from the AnaConDa-S device, as used to deliver inhaled sedation to ARDS patients. The ability to collect fluid from the AnaConDa-S as a surrogate for distal airspace fluid could be important in furthering our understanding of the potential beneficial effects of inhaled sevoflurane in ARDS and for prognostic and predictive enrichment in future ARDS trials.

## Data Availability

All data related to this study protocol will be made available upon request.

